# Executive dysfunction relates to salience network desegregation in behavioral variant frontotemporal dementia

**DOI:** 10.1101/2025.03.04.25323383

**Authors:** Melanie A. Matyi, Hamsanandini Radhakrishnan, Christopher A. Olm, Jeffrey S. Phillips, Philip A. Cook, Emma Rhodes, James C. Gee, David J. Irwin, Corey T. McMillan, Lauren Massimo

## Abstract

**Background:** The organization of the brain into distinct networks increases (i.e., differentiation) during development and decreases (i.e., de-differentiation) during healthy aging, changes that are associated with improvements and worsening of cognition, respectively. Given that behavioral variant frontotemporal degeneration (bvFTD) is a neurodegenerative disease associated with executive dysfunction and selective vulnerability of the salience network, we tested the hypotheses that bvFTD structural networks are de-differentiated compared to cognitively normal controls (CNC) and that network de-differentiation relates to worse executive function.

**Methods:** In a sample of 90 patients with bvFTD and 71 age-matched CNC with diffusion MRI data we generated probabilistic tractography maps and calculated system segregation, a metric that compares within-network to between-network connectivity, to reflect the extent to which brain networks were differentiated. Patients with bvFTD also completed tests of executive function (digit span backwards, phonemic fluency, category fluency) and a control task (lexical retrieval). We assessed group differences in system segregation, reflecting network differentiation, and, within bvFTD, associations between system segregation and neuropsychological test performance.

**Results:** Compared to CNC, patients with bvFTD exhibited lower system segregation of the salience (*p* < .001) and global brain network (*p* = .008). In bvFTD, lower salience network system segregation was associated with worse executive function (*p_corrected_* = .029) but not lexical retrieval.

**Conclusions:** Results demonstrate associations between executive dysfunction and salience network de-differentiation in patients with bvFTD. Our findings indicate that brain network de-differentiation, reflecting reduced neural capacity for specialized processing, may contribute to the emergence of executive dysfunction in bvFTD.

## Introduction

Behavioral variant frontotemporal degeneration (bvFTD) is frequently characterized as a network disorder, with selective vulnerabilities in the salience network early on in the bvFTD disease course, followed by the frontoparietal executive control network (Seeley et al., 2009).

The salience network is instrumental in orchestrating coordinated brain network activation (Menon & Uddin, 2010; Seeley, 2019). For example, salience network coordinates engagement and deactivation of the executive control network to facilitate responses to emotionally relevant stimuli, functions that are clearly linked with socioemotional processes (Seeley, 2019). The majority of prior work examining the clinical consequences of network breakdown in bvFTD have examined behavioral, rather than executive, consequences. Indeed, reduced functional connectivity of the salience network is associated with impairments in social cognition such as apathy (Godefroy et al., 2022), decreased socioemotional sensitivity (Toller et al., 2018) and interpersonal warmth (Toller et al., 2019). Much of this prior work has investigated functional networks, however the role of *structural* network connectivity in bvFTD is less clear.

Structural connectivity reflects the capacity of the network to facilitate functional connectivity, as white matter pathways form the underlying structure that enables, or constrains, dynamic and synchronized functional activity (Baum et al., 2020; Honey et al., 2010). From this view, breakdown of structural connectivity may contribute to the observed breakdown of functional connectivity in bvFTD. Moreover, evidence of structural network breakdown would suggest different, complementary, biological processes at play in the effects of bvFTD on brain organization, as compared to disruption of functional connectivity. Because structural connectivity measures the strength, or probability, of white matter connections between regions, loss of structural connectivity suggests disruptions to myelination or axon development among other cellular mechanisms (Zatorre et al., 2012). On the other hand, functional connectivity measures the extent to which regions have coordinated activity, regardless of structural connections, and thus, loss of functional connectivity may be related to gene expression and cell types (Richiardi et al., 2015; Zhang et al., 2025). Thus, we examined structural brain network changes in bvFTD to provide insights into the potential cellular mechanisms underpinning observed large-scale brain network breakdown in bvFTD, complementary to prior work investigating functional networks.

As reflected in bvFTD criteria (Rascovsky et al., 2011), the core symptoms of bvFTD include behavioral impairment and executive dysfunction. Executive function is related to the dorsolateral prefrontal and cingulate cortices, among other regions (Ardila et al., 2018). Indeed, there is some evidence that executive dysfunction in bvFTD is related to reduced integrity of the cingulum (Tartaglia et al., 2012) and atrophy of related regions, including the cingulate gyrus, inferior and middle frontal cortex, and insula (Lagarde et al., 2013), suggesting there are multiple regions necessary in order to facilitate executive function. These regions associated with executive dysfunction in bvFTD overlap with the salience and frontoparietal control networks (Yeo et al., 2011). Thus, integrity of the salience and frontoparietal control networks may be related to executive dysfunction in bvFTD. To date, the few studies that have examined functional network connectivity in relation to executive function in bvFTD find that poorer performance on executive function tasks are related to lower or disrupted functional connectivity. Indeed, poor verbal fluency is associated with lower integration of frontal cortex functional connectivity and lower global functional connectivity (Filippi et al., 2017; Rittman et al., 2019) and the executive domain of the Frontal Assessment Battery is associated with resting state functional connectivity of the default mode network (Trojsi et al., 2015). This prior work is informative and is indicative of complex relationships between executive dysfunction and brain connectivity, however these studies relied on small samples of patients with bvFTD, functional connectivity, and/or examination of the brain as a whole or of singular regions associated with large-scale brain networks.

Network neuroscience approaches have established that brain networks support complex cognitive functions. Prior work has established that, during development, children have undifferentiated intrinsic networks and that over the course of the lifespan they differentiate in predictable ways forming distinct and differentiated intrinsic brain networks. Brain network differentiation increases in adolescence and then declines in aging (Baum et al., 2017; M. Y. Chan et al., 2014; He et al., 2020). Indeed, there is an emerging literature on the importance of differentiated intrinsic networks to support specialization of cognition (Baum et al., 2017) while at the same time reducing interference among network systems. For example, increasing network differentiation mediates improvements in executive function in youth (Baum et al., 2017).

Conversely, the de-differentiation of networks in healthy aging is associated with poorer episodic memory (M. L. Chan et al., 2021) and de-differentiation of brain networks in patients with Alzheimer’s disease is associated with worse global cognition and memory scores (Ewers et al., 2021). However, network de-differentiation and its consequences on executive dysfunction in bvFTD has not been examined.

The current study examines the cognitive consequences of structural network de- differentiation in bvFTD. To quantify the degree of brain network differentiation, we use system segregation (M. Y. Chan et al., 2014). System segregation reflects the balance of connectivity within a network to connectivity between that network and the rest of the brain network. Thus, highly differentiated networks are reflected by system segregation values greater than zero, indicating that connectivity within networks is greater than connectivity between networks. This metric, system segregation, benefits from a straightforward calculation, facilitating interpretation and decomposition to examine the relative contributions of within and between network connectivity to overall network de-differentiation. Given evidence that bvFTD is selectively vulnerable to salience network disruption (Seeley et al., 2009) and cognition is associated with the frontoparietal control network (Dixon et al., 2018), we hypothesized that the salience and frontoparietal control networks would exhibit de-differentiation in patients with bvFTD compared to cognitively normal controls (CNC) and would predict poorer performance on tests of executive function.

## Methods

### Participants

The Penn Integrated Neurodegenerative Disease Database (INDD) (Toledo et al., 2014) was queried for patients with a clinical diagnosis of probable bvFTD without evidence of primary progressive aphasia (PPA) at the visit when MRI data was acquired with one of the protocols described below and neuropsychological testing within 9 months of MRI acquisition. A total of 90 patients with bvFTD met these criteria and were included in this study. All patients were initially evaluated at the University of Pennsylvania Frontotemporal Degeneration Center by experienced clinicians using published consensus criteria for bvFTD (Rascovsky et al., 2011). Participants and their legal representatives participated in an informed consent procedure approved by the Institutional Review Board at the University of Pennsylvania. All participants had diffusion MRI (dMRI) data that successfully completed preprocessing procedures detailed below. For patients with multiple timepoints (i.e., multiple timepoints with both dMRI and neuropsychological data) meeting these criteria, the session with the lowest mean framewise displacement (FD), a measure of motion during the dMRI scan, was included in analyses. We studied 71 age-matched CNC with a normal cognitive exam and no history or neurologic or psychiatric condition.

### Cognitive assessments

To evaluate the hypothesized role of the salience and frontoparietal control networks in executive dysfunction, we used a composite score of three verbally-mediated measures of executive functioning. We also examined a control measure involving lexical retrieval with minimal executive resource demands (Higby et al., 2019).

The executive function composite was defined as the average z-score of phonemic fluency, category-guided fluency and backward digit span (Aita et al., 2019; Lezak et al., 2012). The two measures of verbal fluency (i.e., phonemic- and category-guided fluency) involve mental search and working memory which are two dimensions of executive function (Park et al., 2022; Rende et al., 2002). Backward digit span requires planning, mental manipulation of information, and working memory, all dimensions of executive function (Groeger et al., 1999; Hester et al., 2004).

To obtain z-scores, norms derived from the National Alzheimer’s Coordinating Center (NACC) Uniform Data Set (UDS) version 3 were applied (Weintraub et al., 2018). These standardized scores facilitated averaging across the 3 executive function tests. For the phonemic fluency test, patients are given 60 seconds to generate as many unique words as possible, excluding proper nouns and numbers, starting with the letter “F”. The score is the total number of correct, non-repeated responses given in 60 seconds. For the category-guided fluency test, patients are given 60 seconds each to generate as many unique words as possible for animals and vegetables. Scores for animals and vegetables were averaged after z-scoring except when only one test was administered. Lastly, for backward digit span, patients are read a series of digits and are then asked to repeat the digits in reverse order. Trials increase in the length of the digit sequence until the patient makes consecutive errors. The score is the digit sequence length of the last successful trial.

We additionally examined performance on a control measure of lexical retrieval. Lexical retrieval is a task involving picture naming that relies largely on semantic knowledge and less on executive resources (Higby et al., 2019). Visual lexical retrieval was assessed with the 30-item version of the Boston Naming Test (BNT) or the Multilingual Naming Test (MINT). Patients are asked to name pictures and are given as much time as possible to respond. NACC UDS version 2 (Weintraub et al., 2009) and version 3 (Weintraub et al., 2018) norms were applied to BNT and MINT scores, respectively, as BNT was administered in UDS2 and MINT was administered in UDS3.

### Neuroimaging Acquisition

Participants underwent MRI an average of 1.4 months (median = 0, same day) with a range of 0 (same day) to 8.9 months from neuropsychological testing. All structural T1 weighted (T1w) MPRAGE MRI were collected with the same protocol, described below. dMRI data was collected across two protocols. Both dMRI protocols collected 30 non-collinear directions at b=1000 s/mm^2^ with at least one b0 volume using single-shot, spin-echo, diffusion-weighted echo planar imaging with fat saturation. All images were acquired on a 3.0 Tesla SIEMENS TIM Trio scanner. Protocol was included as a covariate in all analyses.

T1w data. T1w data was collected using an 8 channel head coil, axial plane with repetition time = 1620 ms, echo time = 3.87 ms, slice thickness = 1.0 mm, flip angle = 15 degrees, matrix = 192 × 256, and in-plane resolution = 0.977 × 0.977 mm.

dMRI data. Data was acquired using one of two protocols. Protocol A: FOV = 245×245; matrix size = 112×112; number of slices = 57; voxel size = 2.1875×2.1875×2.2 mm3; TR = 6700ms; TE = 85ms. Protocol B: FOV = 240x240; matrix size = 128×128; number of slices = 70; voxel size = 1.875×1.875×2.0 mm3; TR = 8100ms; TE = 83ms.

### Neuroimaging Analysis

dMRI data were preprocessed and reconstructed with QSIPrep 0.21.4, which is based on Nipype 1.8.6 (Gorgolewski et al., 2011) (RRID:SCR_002502). Many internal operations of QSIPrep use Nilearn 0.10.1 (Abraham et al., 2014) (RRID:SCR_001362) and Dipy (Garyfallidis et al., 2014). MP-PCA denoising as implemented in MRtrix3’s dwidenoise (Veraart et al., 2016) was applied with a 5-voxel window. Gibbs unringing was then performed using MRtrix3’s mrdegibbs (Kellner et al., 2016). B1 field inhomogeneity was corrected using dwibiascorrect from MRtrix3 with the N4 algorithm (Tustison et al., 2010). FSL’s eddy was used for head motion correction and eddy current correction (Andersson & Sotiropoulos, 2016) with outlier replacement (Andersson et al., 2016). Scans were not acquired with reverse phase-encode blips so synb0 was implemented to generate an undistorted non-diffusion weighted image (Schilling et al., 2019). Outputs of synb0 were fed into FSL’s TOPUP (Andersson et al., 2003) to estimate and remove susceptibility-induced off-resonance fields. The TOPUP-estimated fieldmap was incorporated into the eddy current and head motion correction interpolation. Final interpolation was performed using the jac method. Framewise displacement (FD) using the implementation in Nipype, following the definitions by (Power et al., 2014) were calculated based on the preprocessed DWI. The DWI time-series were resampled to ACPC, generating a preprocessed DWI run in ACPC space with 1.5mm isotropic voxels.

Multi-tissue fiber response functions were estimated using the Dhollander algorithm (Dhollander et al., 2016, 2019). FODs were estimated via constrained spherical deconvolution (CSD) (Tournier et al., 2004, 2008) using an unsupervised multi-tissue method (Dhollander et al., 2016, 2019). A single-shell-optimized multi-tissue CSD (Tournier et al., 2004) was performed using MRtrix3Tissue (https://3Tissue.github.io), a fork of MRtrix3 (Tournier et al., 2019). FODs were intensity-normalized using mtnormalize (Raffelt et al., 2017). Whole-brain probabilistic tractography was performed using the tckgen function with second-order integration over FODs. Streamline count after Spherical-deconvolution Informed Filtering of Tractograms (SIFT) (Smith et al., 2013) was applied to the Schaefer 100 parcellation of the 7-network Yeo intrinsic brain networks (Schaefer 100×7) (Schaefer et al., 2018; Yeo et al., 2011) and used as our measure of structural connectivity. In the Schaefer 100×7 parcellation each region is assigned to one of seven networks, salience (or ventral attention), frontoparietal control, default mode, dorsal attention, visual, somatomotor, and limbic.

To extract cortical thickness measures, T1w data were processed using the Advanced Normalization Tools (ANTs) antsCorticalThickness pipeline (Tustison et al., 2014) (https://github.com/ftdc-picsl/antsct-aging [v0.3.3-p01]). Briefly, intensity nonuniformity was corrected with N4 bias correction (Tustison et al., 2010). Atropos segmentation used template-based priors to divide images into 6 classes (cortical gray matter (GM), subcortical GM, white matter (WM), cerebrospinal fluid (CSF), brainstem, and cerebellum) (Avants et al., 2011). Labels from the Schaefer 100×7 parcellation (Schaefer et al., 2018; Yeo et al., 2011) were transformed from Oasis template space to native space by using the warps made with antsCorticalThickness, and were masked by individual cortex defined using voxel-wise thickness > 0.0001 mm.

### Network differentiation

Network differentiation was measured using system segregation which characterizes differentiation as greater within-network connectivity than between-network connectivity following the equation as previously described (M. Y. Chan et al., 2014):

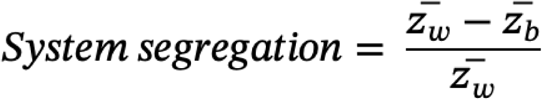

where is the mean connectivity among regions within a given network and is the mean connectivity between regions of a given network and regions of all other networks. Thus, higher system segregation values reflect stronger network differentiation, where within network connectivity is greater than between network connectivity. In contrast, lower system segregation values reflect network de-differentiation, where within-network connectivity is more balanced with, or less than, between-network connectivity. In order to test our hypotheses, we performed statistical tests on system segregation for the salience and frontoparietal control networks, in addition to global segregation, which is the mean segregation of all seven Yeo networks (i.e., salience/ventral attention, frontoparietal control, default mode, dorsal attention, visual, somatomotor, and limbic).

### Statistical Methods

Descriptive statistics were calculated for demographic and clinical variables, using chi-squared tests for categorical variables and t-tests with unequal variances assumption for continuous variables. T-tests were also performed to evaluate gray matter differences between the patient and control groups.

ANCOVA models evaluated the influence of group (patient vs. control) on system segregation of the global, salience, and frontoparietal control networks, adjusted for age at MRI scan, sex, education, mean framewise displacement (FD) during dMRI scan, MRI protocol, and global cortical thickness. Next, linear models evaluated the association of system segregation of global, salience, and frontoparietal control networks on the executive function composite, as well as our control task, lexical retrieval. All models within patients with bvFTD adjusted for disease duration, age at MRI scan, sex, education, dMRI scan mean FD, MRI protocol, and global cortical thickness. Reported p-values were adjusted using false discovery rate (FDR) correction.

We further probed significant effects of system segregation on executive dysfunction by examining the contribution of the components of system segregation (i.e., within and between network connectivity) to determine if the effect of system segregation on executive dysfunction is driven by i) lower within network connectivity and/or ii) higher between network connectivity. Within and between network connectivity were normalized by total network connectivity.

## Results

### Demographic variables

The bvFTD and CNC groups did not differ on age at MRI scan, education, or MRI protocol (see Table 1). The patients with bvFTD had greater motion on dMRI scans (t = 4.17, df = 129.58, *p* < .001) and were more likely to identify as male *χ*^2^ = 12.2, df = 1, *p* < .001), which we covary for in subsequent analyses. We also report means and standard deviations of neuropsychological testing after standardization.

**Table 1.** Sample Characteristics.

|  | bvFTD | Controls | difference |
| --- | --- | --- | --- |
| N | 90 | 71 |  |
| Age, years | 62.9 (7.7) | 61.5 (9.4) | n.s. |
| Motion, frame-wise displacement | .48 (.30) | .33 (.14) | $p < .001$ |
| Education, years | 16.0 (3.1) | 15.9 (2.4) | n.s. |
| Sex (%F) | 34.4% | 63.4% | $p < .001$ |
| dMRI Protocol (A/B) | 18/72 | 14/57 | n.s. |
| Disease duration, years | 4.63 (2.7) | -- |  |
| Animal fluency, Z-score | -1.94 (1.1) | -- |  |
| Vegetable fluency, Z-score | -1.96 (.95) | -- |  |
| Category fluency, Z-score | -1.95 (1.0) | -- |  |
| Phonemic fluency, Z-score | -1.73 (.98) | -- |  |
| Digit span backwards, Z-score | -1.18 (1.2) | -- |  |
| Executive function composite, Z-score | -1.62 (.91) | -- |  |
| Lexical retrieval, Z-score | -1.71 (2.2) | -- |  |

*Note.* Standardized z-scores are reported for all neuropsychological tests. Values in parentheses are standard deviations.

### Cortical atrophy in patients with bvFTD

Patients with bvFTD exhibited greater atrophy in most brain regions, with the greatest degree of atrophy in the frontal and temporal regions as expected. There was one area of the left visual cortex where there was no difference between bvFTD and control participants. Figure 1 illustrates the magnitude of cortical thickness differences between groups for regions with FDR corrected *p* < 0.05 (Mowinckel & Vidal-Piñeiro, 2020).

**Figure 1.**
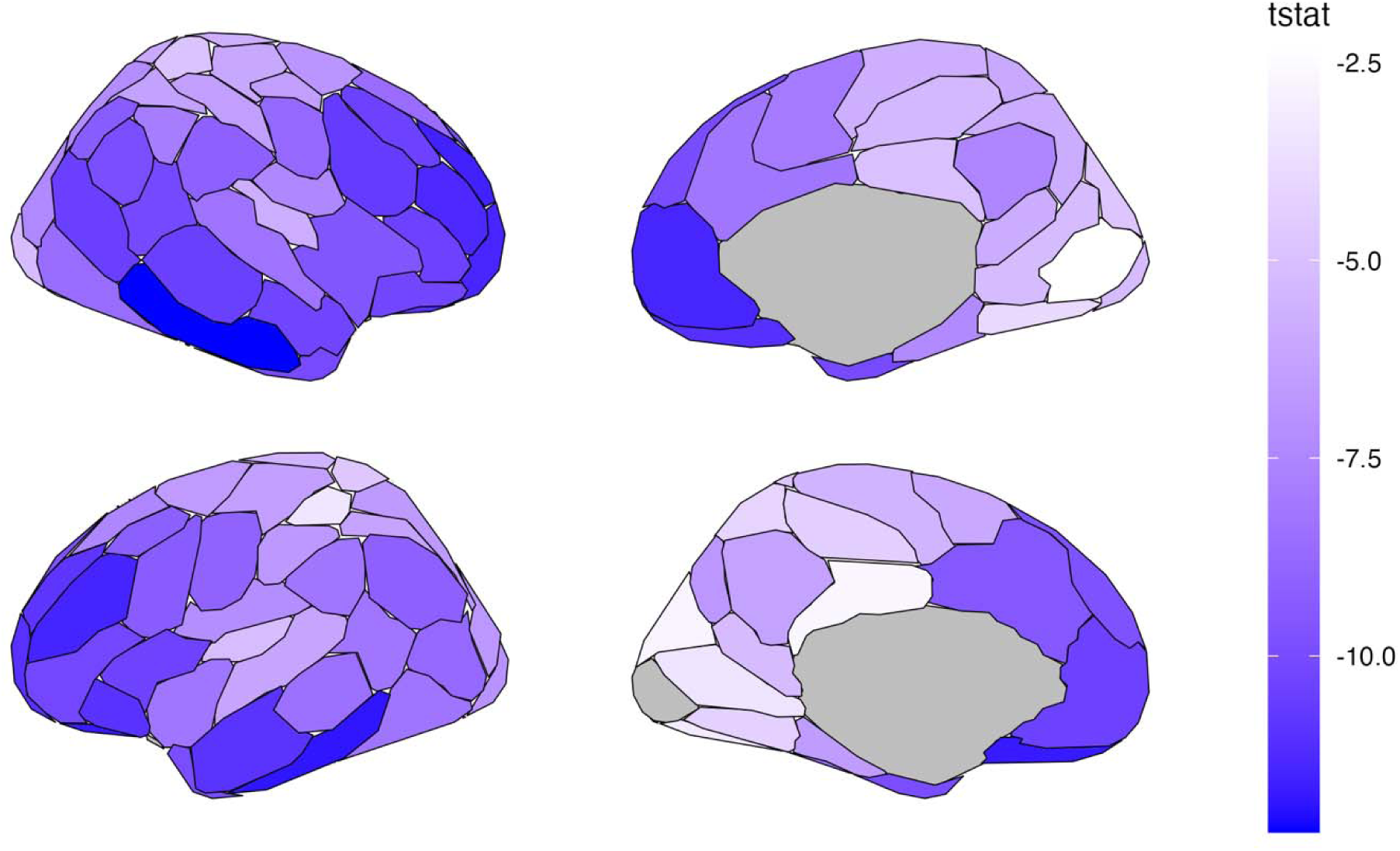
Cortical atrophy in patients with bvFTD compared to CNC. T-statistics are shown for regions with FDR *p_corrected_* < .05.

### De-differentiation of intrinsic brain networks in patients with bvFTD compared to CNC

ANCOVA comparisons evaluated system segregation of the global, salience, and frontoparietal control networks. As illustrated in Figure 2, we observed a significant difference between bvFTD and CNC in global system segregation (F(1, 153) = 7.25, *p* = .008) and salience system segregation (F(1, 153) = 19.32, *p* < .001), where bvFTD has lower system segregation compared to CNC. We did not observe differences in the frontoparietal control network (F(1, 153) = 1.98, *p* = .162).

**Figure 2.**
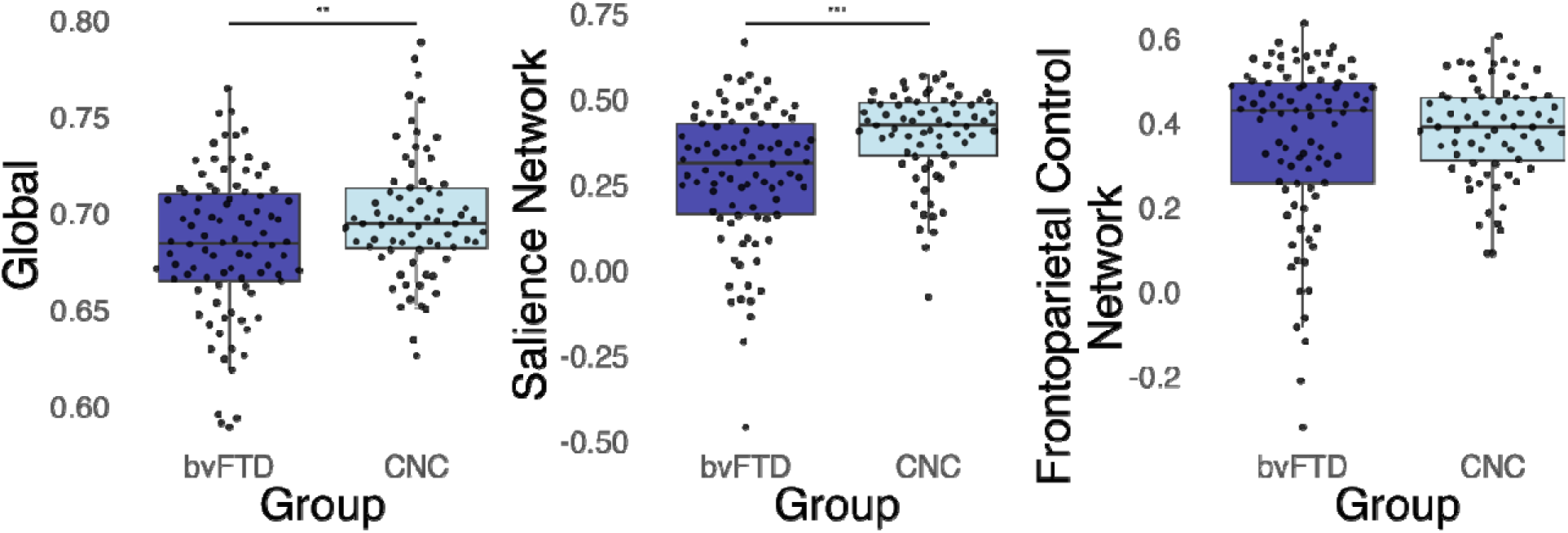
De-differentiation of intrinsic brain networks in patients with bvFTD compared to CNC. The global and salience networks but not frontoparietal control network are de- differentiated in patients with bvFTD compared to CNC. \*\**p* < .01; \*\*\**p* < .001

### De-differentiation of intrinsic brain networks in patients with bvFTD relates to executive dysfunction

Linear regressions evaluated whether system segregation of the global, salience, and frontoparietal control networks was predictive of executive function. System segregation of the salience network was associated with poorer performance across tests of executive functioning (β = 0.26, SE = 0.09, *p_corrected_*= .038). Associations between salience segregation and the individual executive function tests (i.e., phonemic fluency, category-guided fluency and backward digit span) making up the composite score are reported in the supplement (Fig. S1, Table S1). As predicted, we did not observe any associations between system segregation and lexical retrieval (Figure 3). System segregation of global and frontoparietal control networks were not associated with performance on any of the examined neuropsychological tests, suggesting that executive dysfunction is specific to de-differentiation of the salience network.

**Figure 3.**
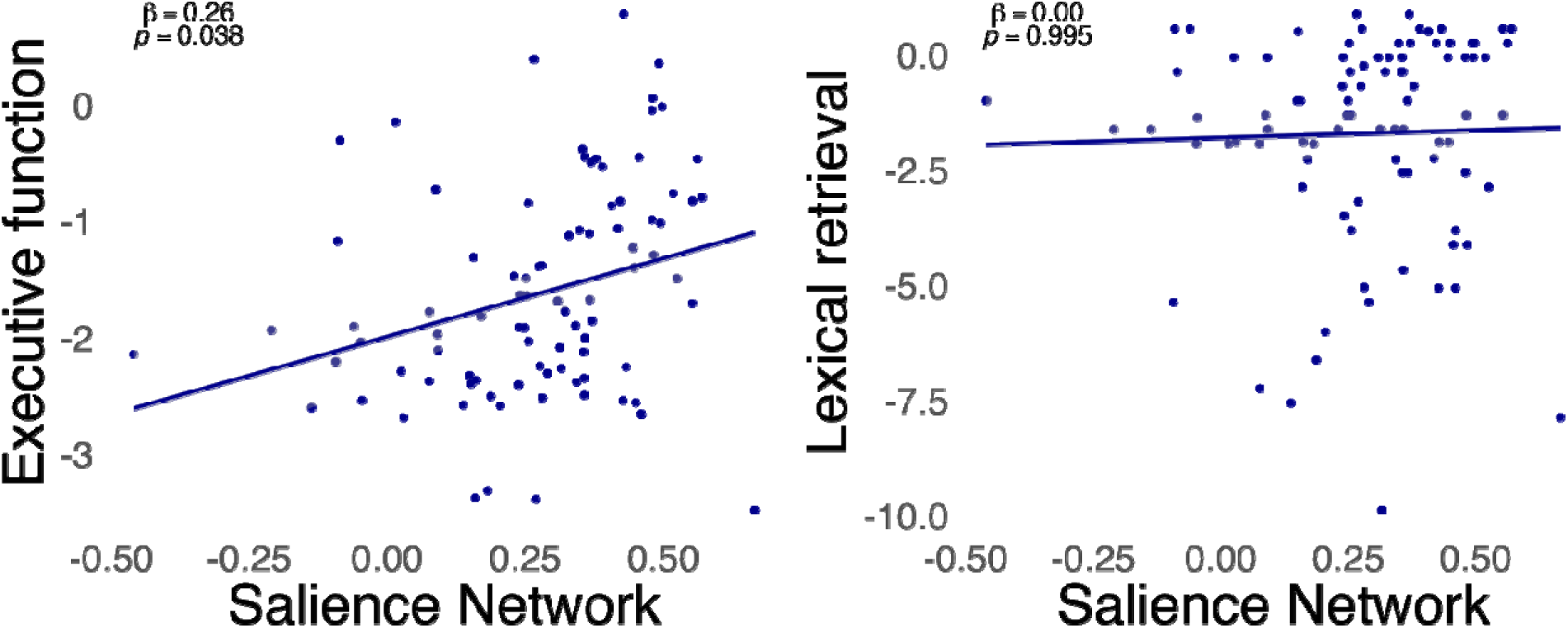
De-differentiation of intrinsic brain networks in patients with bvFTD relates to executive dysfunction. De-differentiation of the salience network, as measured by system segregation, is associated with executive dysfunction but not lexical retrieval in patients with bvFTD. Standardized z-scores of neuropsychological test performance are shown.

**Figure 4.**
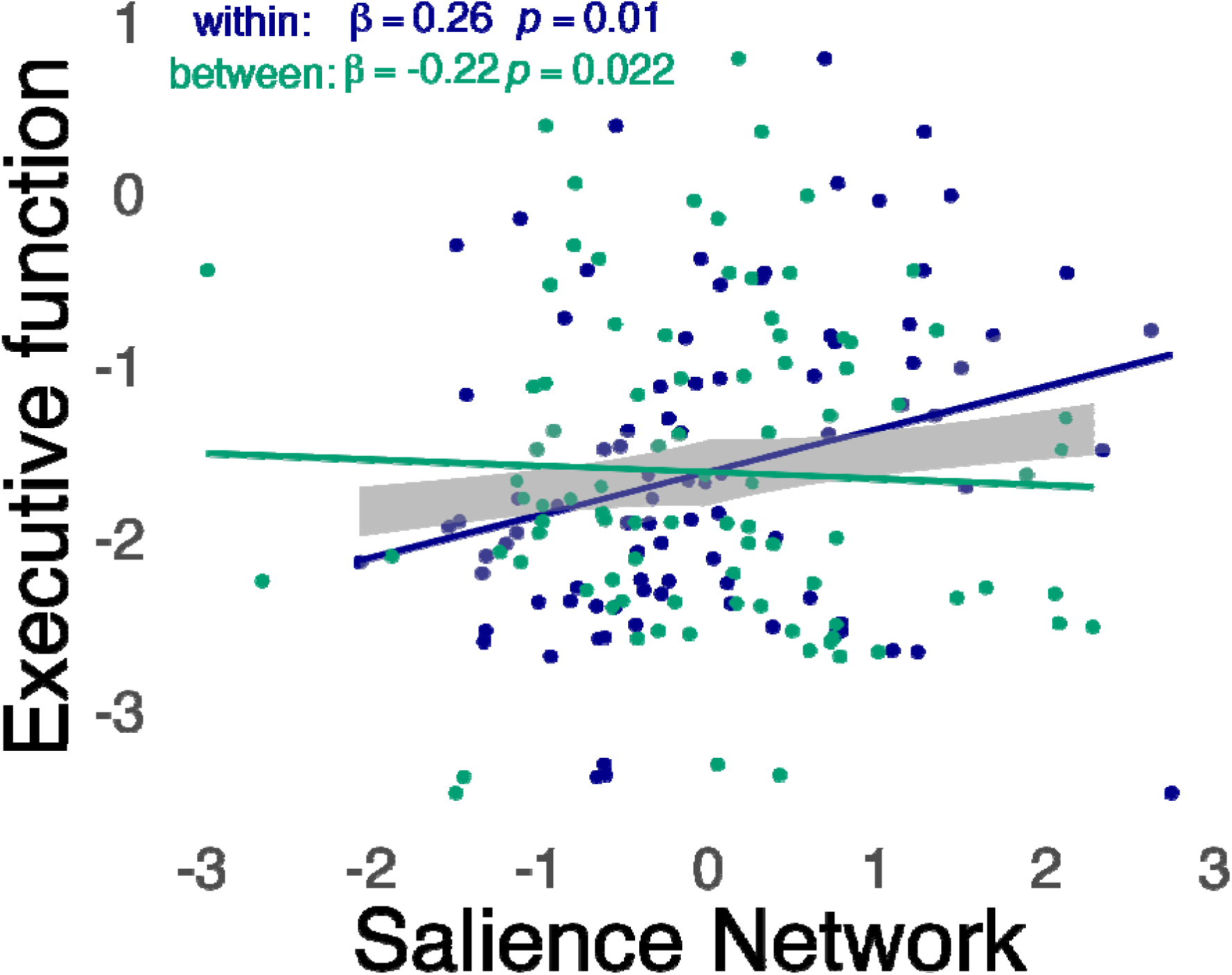
Decomposition of the effects of system segregation on executive dysfunction. Both components of the system segregation in the salience network – lower within network connectivity (dark blue) and higher between network connectivity (green) – are associated with executive dysfunction in patients with bvFTD. Standardized z-scores of neuropsychological test performance are shown. Within and between network connectivity are z-scored for visualization.

### Decomposition of the effects of system segregation on executive dysfunction

To probe our observation that lower system segregation of the salience network is associated with executive dysfunction, we examined the relative contributions of within and between network connectivity of the salience network to executive function. Executive dysfunction was explained both by lower mean connectivity within the salience network (β = .26, SE = 0.10, *p* = .010) and greater mean connectivity between the salience network and non-salience network regions (β = -.22, SE = 0.09, *p* = .022).

## Discussion

In the current study, we examined the contribution of network de-differentiation to executive dysfunction in bvFTD. Results demonstrate that poorer performance across neuropsychological tests with high executive demands (i.e., digit span, phonemic and category fluency) but not low executive demands (i.e., lexical retrieval) are associated with de- differentiation of the salience network as measured by system segregation. Critically, analyses controlled for cortical atrophy, and thus, salience network de-differentiation is a unique contributor to executive dysfunction in bvFTD, above and beyond cortical atrophy. Current findings suggest potential mechanisms for the emergence of executive dysfunction in bvFTD and add insights, complementary to those from functional connectivity studies, into how the bvFTD disease process impacts large scale brain networks.

### De-differentiation of the salience network in patients with bvFTD compared to CNC

Structural brain network de-differentiation reflects the reduced capacity of the brain to support specialized processing. Therefore, our findings suggest that executive dysfunction in bvFTD may emerge, in part, due to a decreasing capacity for the salience network to engage in specialized processing (e.g., ‘bottom-up’ attention to behaviorally relevant stimuli) (Corbetta & Shulman, 2002). Moreover, our observation of de-differentiation of structural brain networks suggests potential cellular mechanisms, distinct from those which follow from prior work examining functional brain networks (Ferreira et al., 2022; Zhang et al., 2025). Structural connectivity measures the strength, or probability, of white matter connections between regions. In turn, loss of structural connectivity suggests disruptions to myelination (e.g., thickness, morphology), axon development (e.g., branching, sprouting, packing density, diameter, number) and/or astrocyte morphology or number, among other cellular mechanisms (Zatorre et al., 2012).

Importantly, patients with bvFTD, compared to CNC, exhibited significant salience network de- differentiation in addition to cortical thinning, indicating that effects of network de- differentiation are not attributable to overall cortical atrophy. Thus, our findings provide novel insights into potential white matter mechanisms of neurodegeneration in bvFTD.

Much of the prior research examining the re-organization and degradation of brain networks in bvFTD have leveraged functional MRI and functional connectivity analysis (Ferreira et al., 2022). Here, we leveraged structural connectivity to examine how the capacity of brain networks to facilitate coordinated activity supports executive functioning. Present findings, derived from diffusion MRI, demonstrate that breakdown in network configurations, analogous to those observed in functional networks, exist at the structural level. Evidence of the breakdown of structural networks underlying functional networks has implications for how functional connectivity findings are interpreted. Together, prior functional, and current structural, connectivity findings suggest that structural network breakdown impacts the ability of functional networks to be recruited. Future work should examine coupling of dMRI with fMRI to assess the extent to which structural network de-differentiation explains functional network dynamics, particularly in the context of executive dysfunction and neuropsychiatric symptoms in bvFTD.

### De-differentiation of the salience network in patients with bvFTD relates to executive dysfunction

Expanding upon prior work, our findings demonstrate the executive dysfunction consequences related to salience network de-differentiation as measured by system segregation. In examining the components of system segregation, we observed that each component significantly contributed to lower executive functioning in patients with bvFTD: both lower connectivity within the salience network and greater connectivity between the salience network and all other networks were related to executive dysfunction, with the within network effect being the largest. This pattern of findings indicates that de-differentiation, or breakdown, of the salience network is associated with executive dysfunction and that this association is primarily driven by the loss of white matter connections among regions that are expected to have high interconnectivity, with a smaller contribution of white matter connections facilitating integration across networks. In other words, executive dysfunction is related to a loss of connections within the salience network with additional contribution of a reduced capacity for the salience network to interact with other brain networks.

Our observation of significant contributions of structural brain network de-differentiation to executive dysfunction in patients with bvFTD suggests a possible mechanism for prior evidence of associations between functional connectivity dynamics and cognitive functioning.

Dynamic *functional* connectivity analyses, which examine functional connectivity patterns over time, indicate that dynamic segregation (i.e., high within network connectivity of functional networks) and integration (i.e., high connectivity between functional networks), and the ability to switch between these states is associated with cognitive abilities, as measured by a cognitive composite score and, in particular, processing speed (Wang et al., 2021). Specifically, longer switch times between ‘segregated’ and ‘integrated’ states, and less time spent in a segregated state are associated with poorer performance on executive tasks. One possibility for longer switch times between segregated and integrated states may be the breakdown of salience network structure which, in turn, may prevent the salience network from facilitating efficient switching of states. Moreover, the salience, or ventral attention, network is critical to stimulus-driven, or bottom-up, attention (Corbetta & Shulman, 2002). In this role, the salience network directs attention towards salient, behaviorally relevant stimuli by interacting with other brain networks. Therefore, a breakdown in salience network structure may result in a downstream impact of poor recruitment of other functional networks, together resulting in executive dysfunction. Indeed, results from prior work, utilizing functional connectivity, suggests that cognitive decline is related to less selective recruitment of brain networks during cognitive tasks (He et al., 2020). The identification of similar patterns of network de-differentiation at the level of the structural brain network in the present study, as have previously been observed with functional networks, adds to current network hypotheses of executive dysfunction in bvFTD. Our findings further suggest that executive dysfunction is related to the breakdown of the brain structure supporting functional networks, beyond cortical atrophy.

The observed relationship between executive dysfunction and salience network de- differentiation may be related to the critical role the salience network plays in facilitating coordinated activation and deactivation of other large-scale networks, including the frontoparietal control network (Goulden et al., 2014). Our results, highlighting the role of de- differentiation of salience, but not frontoparietal control network, suggest that the executive dysfunction observed in bvFTD may result from the lack of recruitment, rather than direct dysfunction, of the frontoparietal control network by the salience network. In other words, we suggest that *structural* de-differentiation of the salience network may attenuate the capacity of the network to facilitate recruitment of the frontoparietal control network in response to task demands, resulting in poor performance on tasks requiring high executive demand. That is, if structural connectivity constrains/facilitates functional connectivity, then de-differentiation of the salience network’s structure would likely contribute to disrupted functional network organization. Future work should investigate this by assessing the interaction of structural and functional network dynamics between the intrinsic networks.

The present study demonstrates that structural network de-differentiation in bvFTD has consequences for executive dysfunction. This adds to extant research which has focused on network contributions to socioemotional dysfunction in bvFTD. Critically, our finding of the selective impact of salience network de-differentiation on executive dysfunction indicate that the consequences of salience network de-differentiation extend beyond socioemotional processes.

Furthermore, salience network de-differentiation contributed to executive dysfunction above and beyond whole brain cortical atrophy, indicating that desegregation explains additional variance in executive dysfunction beyond cortical atrophy. In other words, preserved salience network structure may attenuate the effect of cortical atrophy on executive dysfunction, similar to findings in Alzheimer’s disease (Ewers et al., 2021). The links between salience network integrity and executive function demonstrated here should be further explored, as salience network de-differentiation may represent a mechanism for cognitive decline in bvFTD. Alternatively, preserved structural organization, that is maintenance of network differentiation, of the salience network in patients with bvFTD may represent resilience to pathology.

Strengths of this study include inclusion of cortical atrophy in our models. Although atrophy explains a portion of variance in executive function, salience desegregation remains a significant predictor after controlling for atrophy. Furthermore, we demonstrate specificity in our findings. Salience de-differentiation, but not global de-differentiation, was related to executive dysfunction, and was not related to performance on a lexical retrieval task. In an additional effort to demonstrate the specificity of salience de-differentiation to executive dysfunction, we excluded patients with bvFTD and symptoms of primary progressive aphasia to reduce the potential impact of language impairments on findings.

The strengths of this study should be considered in light of some limitations. First, although the study benefits from a large sample of patients with bvFTD, analyses were based on structural networks calculated from 30 direction dMRI data, and thus our data are limited by low angular resolution. Second, MRI data was pooled across highly similar, but different protocols. Third, our patient sample was characterized only by clinical phenotype; different underlying proteinopathies and biofluid biomarkers were not considered. bvFTD is pathologically heterogeneous with FTLD-tau in 40% of cases and FTLD-TDP in 50% of cases (Bahia et al., 2013). Exploring differences in network segregation by underlying proteinopathy would be an interesting future direction as evidence demonstrates convergence of tau and TDP-43 pathology on select paralimbic cells in the salience network (Perry et al., 2017; Seeley et al., 2006, 2008, 2009) but partially distinct cellular degeneration patterns (Chen et al., 2022; Irwin et al., 2018; Ohm et al., 2025). Additionally, our sample is highly homogeneous in regards to demographic makeup, with the majority of participants identifying as white and highly educated. Lastly, this study utilized a cross-sectional design, and thus, we are unable to make any conclusions about the direction of effect between network desegregation and executive function deficits.

In summary, structural network de-differentiation may represent a mechanism of executive dysfunction in bvFTD. Alternatively, preserved salience network de-differentiation, despite cortical atrophy, may reflect resilience to cognitive decline in bvFTD. Future work should examine the association of executive dysfunction and network de-differentiation longitudinally to assess whether network de-differentiation predicts executive dysfunction and to tease out structure-function network dynamics that may contribute to cognitive decline. The identification of biological mechanisms underlying the process of network de-differentiation will provide more insights to inform symptom management in bvFTD by suggesting modifiable mechanisms to target. Furthermore, network de-differentiation in bvFTD may also reflect cognitive reserve, suggesting mechanisms of cognitive resilience in bvFTD. In Alzheimer’s disease, evidence of preserved segregation as a reflection of cognitive resilience exists: at similar disease severity, patients with greater network differentiation had better than expected cognition (Ewers et al., 2021). Future work should examine interactions of disease stage and network desegregation in relation to executive dysfunction in bvFTD.

## Conclusion

We demonstrate that the structural integrity of the salience network is compromised in patients with bvFTD compared to control participants, in line with prior findings highlighting salience network dysfunction in bvFTD. We further show the executive consequences of structural de-differentiation of the salience network in patients with bvFTD. Reduced differentiation, as measured by lower segregation, of the salience network in patients with bvFTD, relates to poorer performance on neuropsychological evaluation of executive function. Because structural brain network de-differentiation reflects the reduced capacity of the brain network to support specialized processing, current findings suggest a potential mechanism for the emergence of executive dysfunction in bvFTD. Critically, all analyses controlled for cortical atrophy, and thus, salience network de-differentiation is a unique contributor to executive dysfunction in bvFTD. This assessment of structural network organization provides insight into the brain architecture that contributes to executive dysfunction in bvFTD; structural brain network de-differentiation, reflecting reduced neural capacity for specialized processing, may mediate emergence of executive dysfunction in bvFTD.

## Data Availability

Data will be made available upon reasonable request

## Funding

This work was supported by NIH funding (P01AG066597, T32AG076411, P30AG072979, R01AG076832, R01AG090414, R01AG054519), Penn Institute on Aging, and DeCrane Fund for Primary Progressive Aphasia.

## Author Contributions

Melanie A. Matyi: Conceptualization, Formal analysis, Writing – Original Draft, Visualization; Hamsanandini Radhakrishnan: Software, Data Curation, Writing – Review & Editing; Christopher A. Olm: Software, Data Curation, Writing – Review & Editing; Jeffrey S. Phillips: Resources, Writing – Review & Editing, Funding acquisition; Philip A. Cook: Software, Data Curation, Writing – Review & Editing; Emma Rhodes: Writing – Review & Editing; James C. Gee: Resources, Funding acquisition; David J. Irwin: Resources, Writing – Review & Editing, Funding acquisition; Corey T. McMillan: Conceptualization, Resources, Writing – Review & Editing, Funding acquisition, Supervision; Lauren Massimo: Conceptualization, Resources, Writing – Review & Editing, Funding acquisition, Supervision

## Notes

### Competing Interest Statement

The authors have declared no competing interest.

### Author Declarations

The IRB of the University of Pennsylvania gave ethical approval for this work.

### Summary of Updates

Decomposition analysis added. Revisions throughout manuscript for clarity.

