## Supplement for "Executive dysfunction relates to salience network desegregation in behavioral variant frontotemporal dementia"

|  | β | SE | *p* (uncorrected) |
| --- | --- | --- | --- |
| Phonemic fluency | 0.21 | 0.10 | .033 |
| Category fluency | 0.23 | 0.09 | .020 |
| Digits backward | 0.23 | 0.10 | .028 |

Table S1. Association of executive functioning measures with salience network segregation.

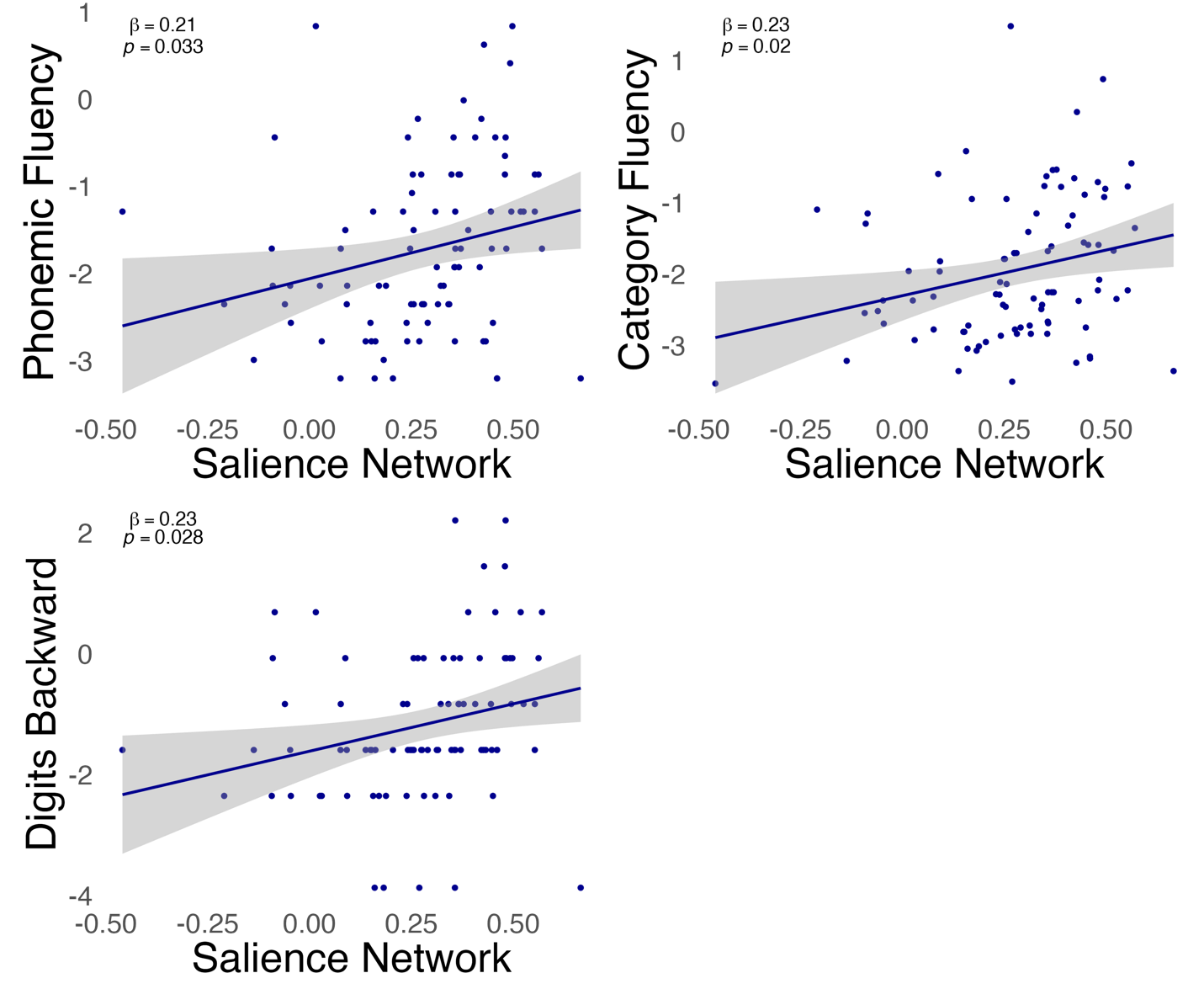

Figure S1. De-differentiation of the salience network in patients with bvFTD relates to executive dysfunction. Lower salience network system segregation is associated with executive dysfunction as measured by phonemic fluency, category-guided fluency and digit span backwards.
